# Alpha-1 blockers and susceptibility to COVID-19 in benign prostate hyperplasia patients : an international cohort study

**DOI:** 10.1101/2021.03.18.21253778

**Authors:** Akihiko Nishimura, Junqing Xie, Kristin Kostka, Talita Duarte-Salles, Sergio Fernández Bertolín, María Aragón, Clair Blacketer, Azza Shoaibi, Scott L DuVall, Kristine Lynch, Michael E Matheny, Thomas Falconer, Daniel R Morales, Mitchell M Conover, Seng Chan You, Nicole Pratt, James Weaver, Anthony G Sena, Martijn J Schuemie, Jenna Reps, Christian Reich, Peter R Rijnbeek, Patrick B Ryan, George Hripcsak, Daniel Prieto-Alhambra, Marc A Suchard

**Author notes:** **Corresponding author** Prof Daniel Prieto-Alhambra, Centre for Statistics in Medicine, Botnar Research Centre, Windmill Road, OX37LD, Oxford, United Kingdom.

## Abstract

Alpha-1 blockers, often used to treat benign prostate hyperplasia (BPH), have been hypothesized to prevent COVID-19 complications by minimising cytokine storms release. We conducted a prevalent-user active-comparator cohort study to assess association between alpha-1 blocker use and risks of three COVID-19 outcomes: diagnosis, hospitalization, and hospitalization requiring intensive services. Our study included 2.6 and 0.46 million users of alpha-1 blockers and of alternative BPH therapy during the period between November 2019 and January 2020, found in electronic health records from Spain (SIDIAP) and the United States (Department of Veterans Affairs, Columbia University Irving Medical Center, IQVIA OpenClaims, Optum DOD, Optum EHR). We estimated hazard ratios using state-of-the-art techniques to minimize potential confounding, including large-scale propensity score matching/stratification and negative control calibration. We found no differential risk for any of COVID-19 outcome, pointing to the need for further research on potential COVID-19 therapies.

## Introduction

As the number of infections with severe acute respiratory syndrome coronavirus 2 (SARS-CoV-2) continues to increase, so does the search for therapies to prevent its respiratory and multi-organ complications. Treatments under study have been classified into antiviral/repurposed ones, aiming to reduce viral uptake, and concomitant/adjunctive ones, aiming to minimise the risk of or to treat complications (Sanders et al. 2020). Despite the large number of proposed therapies, most drug trials have been negative: an increasing list of therapies including azithromycin, hydroxychloroquine, lopinavir/ritonavir, and tocilizumab have little or no impact on mortality or patient-relevant outcomes; whilst only corticosteroids and remdesivir have potential effects on mortality, mechanical ventilation, length of hospital stay, or duration of symptoms (Siemieniuk et al. 2020). Given the scarcity of available treatments, the search for medicines with putative effects to treat COVID-19 and minimize its complications is due to continue.

Activation of inflammatory and related cascades are part of innate immunity and crucial in the immune response against SARS-CoV-2. However, aggressive inflammatory response to SARS-CoV-2, known as cytokine release syndrome (CRS), has been recognized as a driving cause of high morbidity and mortality in COVID-19. In animal studies using mice, a direct disruption of catecholamine synthesis reduced cytokine release and offered protection against lethal complications of CRS (Staedtke et al. 2018). The same study found a similar protection from use of prazosin, an alpha-1 adrenergic receptor antagonist (alpha-1 blocker), to block catecholamines signalling. It has therefore been postulated that CRS may be prevented by targeting the catecholamine-cytokine axis. Vogelstein et al. 2020 investigated this hypothesis through a cohort study among patients with pneumonia and acute respiratory distress, in which alpha-1 blocker users were found to have a lower risk of mechanical ventilation and death compared to non-users. However, the interplay between catecholamines and immune-inflammatory regulation is complex and dynamic; the net effect of inhibition of catecholamines on COVID-19 thus remains uncertain (Gubbi et al. 2020).

While at least one randomized controlled trial is ongoing to study the efficacy of prazosin for SARS-CoV-2 infection, no robust evidence has yet been reported regarding the effect of alpha-1 blockers on either the prevention or the treatment of COVID-19. In this article, therefore, we investigated the association between the use of alpha-blockers and the susceptibility to COVID-19 disease (diagnosis), and related hospitalization and requirement of intensive services.

## Results

### Baseline characteristics

We found in total 2,628,170 users of alpha-1 blockers and 464,525 users of alternative BPH therapy — 5-alpha reductase inhibitors and phosphodiesterase type 5 inhibitors (5ARI/PDE5) — with diagnosis of BPH, all of whom were included in our propensity score (PS) stratified analyses. For PS matched analysis, 2,426,765 (92.3%) and 463,113 (99.7%) of the subjects could be matched based on their baseline characteristics. Table 1 summarizes key socio-demographics, medical history and medicine use for alpha-1 blocker and active comparator users before and after PS stratification and matching. Table 1 focuses on OpenClaims and a selected subset of clinical covariates for an illustrative purpose, but Supplementary Table 1 provides detailed baseline characteristics for all participants in the six contributing data sources. On average, users of alpha-1 blockers were younger and healthier than users of active comparator medicines. For the most part, PS matching and stratification successfully reduced the differences in baseline characteristics to the negligible level of standardized mean differences (SMD) below 0.1 (Figure 2). Due to the relatively small cohort size, PS matching and stratification were less successful in CUIMC. A few covariates remained imbalanced (SMD > 0.1) after PS stratification in SIDIAP and Optum EHR. Except for these cases, no substantial differences in baseline characteristics remained after PS matching and stratification.

**Table 1:** Baseline patient characteristics for alpha-1 blocker and 5ARI/PDE5 user cohorts in the OpenClaims data source. For each target (T) and comparator (C) cohort, we report the proportion of initiators satisfying selected base-line characteristics and the standardized difference of population proportions (SMD) before and after stratification. The smaller SMDs after propensity score adjustment demonstrates improved balance between the two cohorts.

| Characteristic | OpenClaims |  |  |  |  |  |
| --- | --- | --- | --- | --- | --- | --- |
|  | Before stratification |  |  | After stratification |  |  |
|  | *N* = 1,995,594 (T) and 366,734 (C) |  |  |  |  |  |
|  | T (%) | C (%) | SMD | T (%) | C (%) | SMD |
| Age group |  |  |  |  |  |  |
| <25 | 0.1 | 0.1 | 0.00 | 0.1 | 0.1 | 0.00 |
| 25-29 | 0.1 | 0.1 | 0.00 | 0.1 | 0.1 | 0.00 |
| 30-34 | 0.1 | 0.1 | -0.01 | 0.1 | 0.1 | -0.01 |
| 35-39 | 0.2 | 0.2 | -0.01 | 0.2 | 0.2 | 0.00 |
| 40-44 | 0.4 | 0.4 | 0.01 | 0.4 | 0.4 | 0.01 |
| 45-49 | 1.2 | 0.8 | 0.04 | 1.2 | 1.0 | 0.01 |
| 50-54 | 3.1 | 1.8 | 0.08 | 2.9 | 2.7 | 0.01 |
| 55-59 | 7.0 | 4.1 | 0.13 | 6.6 | 6.2 | 0.02 |
| 60-64 | 12.2 | 8.1 | 0.14 | 11.6 | 11.3 | 0.01 |
| 65-69 | 17.4 | 13.9 | 0.10 | 16.8 | 16.9 | 0.00 |
| 70-74 | 19.2 | 19.2 | 0.00 | 19.2 | 19.1 | 0.00 |
| 75-79 | 16.7 | 19.2 | -0.07 | 17.1 | 17.4 | -0.01 |
| 80-84 | 17.1 | 24.1 | -0.18 | 18.1 | 18.7 | -0.02 |
| 85-89 | 5.3 | 7.8 | -0.10 | 5.7 | 5.8 | 0.00 |
| 90-94 |  |  |  |  |  |  |
| 95+ |  |  |  |  |  |  |
| Medical history: General |  |  |  |  |  |  |
| Chronic liver disease | 0.7 | 0.4 | 0.05 | 0.7 | 0.6 | 0.01 |
| Chronic obstructive lung disease | 7.0 | 5.4 | 0.07 | 6.8 | 6.7 | 0.00 |
| Dementia | 1.8 | 2.3 | -0.03 | 1.9 | 1.9 | 0.00 |
| Diabetes mellitus | 19.3 | 16.0 | 0.09 | 18.8 | 18.6 | 0.00 |
| Hyperlipidemia | 29.8 | 28.9 | 0.02 | 29.7 | 29.6 | 0.00 |
| Hypertensive disorder | 38.2 | 34.8 | 0.07 | 37.7 | 37.6 | 0.00 |
| Obesity | 3.9 | 2.7 | 0.07 | 3.7 | 3.5 | 0.01 |
| Renal impairment | 11.2 | 9.7 | 0.05 | 11.1 | 10.8 | 0.01 |
| Medical history: Cardiovascular disease |  |  |  |  |  |  |
| Cerebrovascular disease | 3.3 | 3.2 | 0.00 | 3.3 | 3.3 | 0.00 |
| Ischemic heart disease | 4.0 | 3.6 | 0.03 | 4.0 | 4.0 | 0.00 |
| Medical history: Neoplasms |  |  |  |  |  |  |
| Malignant neoplastic disease | 11.1 | 9.7 | 0.04 | 10.9 | 10.7 | 0.01 |
| Primary malignant neoplasm of prostate | 4.8 | 2.9 | 0.10 | 4.6 | 4.2 | 0.02 |
| Medication use |  |  |  |  |  |  |
| Antiinflammatory and antirheumatic products | 26.0 | 19.6 | 0.15 | 25.0 | 24.6 | 0.01 |
| Antineoplastic agents | 5.5 | 5.4 | 0.00 | 5.5 | 5.6 | 0.00 |
| Antithrombotic agents | 25.5 | 24.8 | 0.01 | 25.4 | 25.5 | 0.00 |
| Drugs used in diabetes | 26.1 | 21.5 | 0.11 | 25.5 | 25.4 | 0.00 |
| Immunosuppressants | 2.8 | 2.3 | 0.03 | 2.8 | 2.7 | 0.00 |

**Figure 2.**
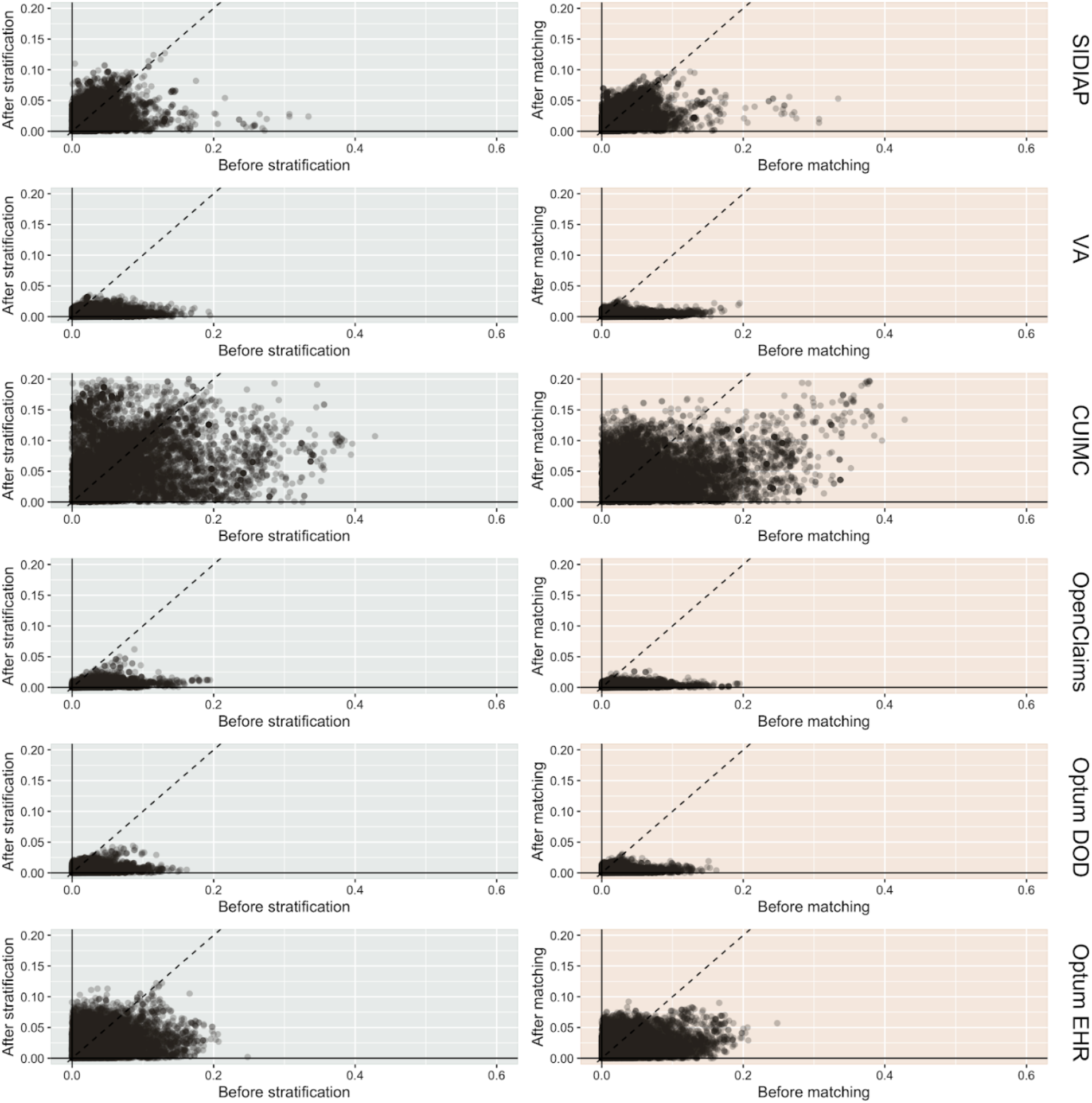
Cohort balance diagnostics comparing alpha-1 blocker and 5ARI/PDE5 prevalent users. We plot the absolute SMD of population proportions for all available patient characteristics (13,950 in SIDIAP, 81,436 in VA, 24,807 in CUIMC, 73,113 in OpenClaims, 79,184, in Optum DOD, 40,621 in Optum EHR) before and after propensity score stratification or matching across data sources. CUIMC fails study diagnostics under both stratification and matching since the absolute SMDs are not consistently < 0.1. SIDIAP and Optum EHR fail study diagnostics under stratification only.

### Incidence rates of COVID-19-related outcomes

We report in Table 2 patient cohort size for each data source, in PS stratified and PS matched, and number of participants with each of the three outcomes of interest. In the PS-matched cohorts (excluding CUIMC), in total 6,319 alpha-1 blocker users of (275 in SIDIAP, 1,485 in VA, 4,351 in OpenClaims, 175 in Optum DOD and 33 in Optum EHR) and 1,105 5ARI/PDE5 users (51 in SIDIAP, 236 in VA, 764 in OpenClaims, 47 in Optum DOD, and 7 in Optum EHR) were diagnosed with COVID-19. Incidence rates of COVID-19 diagnosis were 85.63/1,000 person-years amongst alpha-1 blockers, and 108.25/1,000 among 5ARI/PDE5 users in SIDIAP; 8.96 vs 7.97/1,000 in VA; 5.62 vs 4.78/1,000 in OpenClaims; 3.40 vs 4.89/1,000 in Optum DOD; and 38.88 vs 47.08/1,000 in Optum EHR. Similarly, a total of 3,108 alpha-1 blockers and 563 5ARI/PDE5 users were hospitalized with COVID-19. Incidence rates of hospital admission ranged from 2.29/1,000 in alpha-1 blocker users and 3.64/1,000 in 5ARI/PDE5 users in Optum DOD to 35.66 vs 42.2/1,000 in SIDIAP. Finally, 110 (92 in VA, 18 in Optum DOD) alpha-1 blocker users and 18 (12 in VA, 6 in Optum DOD) 5ARI/PDE5 users received intensive services, with incidence rates of 0.55 vs 0.40/1,000 respectively in VA, and 0.35 vs 0.62/1,000 in Optum DOD.

**Table 2:** Populations and COVID-19 outcomes for alpha-1 blocker (T) and 5ARI/PDE5 (C) user cohorts. We report population size, total exposure time, outcome events (Covid diagnosis, hospitalization, and intensive services) and minimally detectable risk ratio (MDRR). MDRR is provided only for Covid diagnosis due to the space constraint. The database abbreviations are defined under the heading Data Sources in the Method section.

|  | Patients |  | Time (years) |  | Diagnosis |  | Hospital |  | Intensive |  | MDRR<br>(Diagnosis) |
| --- | --- | --- | --- | --- | --- | --- | --- | --- | --- | --- | --- |
|  | T | C | T | C | T | C | T | C | T | C |  |
| Stratified analysis |  |  |  |  |  |  |  |  |  |  |  |
| SIDIAP | 11,793 | 1,318 | 4,162 | 471 | 334 | 51 | 132 | 20 | 0 | 0 | 1.61 |
| VA | 360,802 | 54,723 | 189,564 | 29,642 | 1,854 | 236 | 636 | 96 | 111 | 12 | 1.20 |
| CUIMC | 2,414 | 582 | 338 | 84 | 27 | <5 | 16 | <5 | 0 | 0 | 4.53 |
| OpenClaims | 1,995,594 | 366,734 | 817,994 | 160,225 | 4,809 | 767 | 2,621 | 407 | 0 | 0 | 1.11 |
| Optum DOD | 241,842 | 39,032 | 56,438 | 9,613 | 193 | 47 | 131 | 35 | 18 | 6 | 1.69 |
| Optum EHR | 15,275 | 2,136 | 1,031 | 149 | 50 | 7 | 32 | 5 | <5 | 0 | 3.10 |
| Matched analysis |  |  |  |  |  |  |  |  |  |  |  |
| SIDIAP | 8,994 | 1,315 | 3,211 | 471 | 275 | 51 | 115 | 20 | 0 | 0 | 1.59 |
| VA | 312,522 | 54,642 | 165,688 | 29,600 | 1,485 | 236 | 495 | 96 | 92 | 12 | 1.21 |
| CUIMC | 1,873 | 520 | 261 | 74 | 18 | <5 | 11 | <5 | 0 | 0 | 6.58 |
| OpenClaims | 1,873,014 | 365,534 | 774,635 | 159,742 | 4,351 | 764 | 2,361 | 407 | 0 | 0 | 1.11 |
| Optum DOD | 218,032 | 38,988 | 51,451 | 9,602 | 175 | 47 | 118 | 35 | 18 | 6 | 1.69 |
| Optum EHR | 12,303 | 2,114 | 848 | 148 | 33 | 7 | 19 | 5 | <5 | 0 | 3.50 |

### Hazard ratios of COVID-19-related outcomes

Table 3 and Figure 3 present database-specific calibrated HRs for the risk of COVID-19 diagnosis, hospitalization, and internsive services under both PS stratified and matched analyses. Findings from both analyses are consistent with each other, but here we focus on the PS matched cases that passed the proposed diagnostics, which included all data sources except CUIMC. The risk of COVID-19 diagnosis did not differ between alpha-1 blocker and 5ARI/PDE5 users in any of the data sources, with PS-matched calibrated HR of 0.99 (95%CI 0.71 - 1.36) in SIDIAP, 1.03 (0.83 - 1.28) in VA, 1.04 (0.90 - 1.21) in OpenClaims, 0.75 (0.51 - 1.11) in Optum DOD, and 1.79 (0.46 - 6.92) in Optum EHR. The meta-analysis yields calibrated HR of 1.02 (95%CI 0.92 - 1.13) for COVID-19 diagnosis.

**Table 3.** Hazard ratios of COVID-19 diagnosis, hospitalization, and intensive services for alpha-1 blocker and 5ARI/PDE5 prevalent-use. We report calibrated hazard ratios (HRs) and their 95% confidence intervals (CIs) and calibrated p-value (*p*), with PS stratification or matching and across data sources. Grayed out entries do not pass study diagnostics and are excluded from the meta-analysis.

|  |  | PS-stratified |  |  | PS-matched |  |  |
| --- | --- | --- | --- | --- | --- | --- | --- |
| | | HR | 95% CI | $p$ | HR | 95% CI | $p$ |
| Diagnosis |  |  |  |  |  |  |  |
|  | SIDIAP | 1.13 | (0.84 - 1.53) | 0.54 | 0.99 | (0.71 - 1.36) | 0.64 |
|  | VA | 1.02 | (0.83 - 1.26) | 0.81 | 1.03 | (0.83 - 1.28) | 0.76 |
|  | CUIMC | 2.54 | (0.80 - 8.01) | 0.14 | 3.65 | (0.67 - 19.9) | 0.15 |
|  | OpenClaims | 1.04 | (0.90 - 1.22) | 0.58 | 1.04 | (0.90 - 1.21) | 0.56 |
|  | Optum DOD | 0.69 | (0.49 - 0.97) | 0.03 | 0.75 | (0.51 - 1.11) | 0.15 |
|  | Optum EHR | 1.46 | (0.55 - 3.85) | 0.43 | 1.79 | (0.46 - 6.92) | 0.39 |
|  | Meta-analysis | 1.03 | (0.94 - 1.12) | 0.54 | 1.02 | (0.92 - 1.13) | 0.68 |
| + Hospitalization |  |  |  |  |  |  |  |
|  | SIDIAP | 1.26 | (0.78 - 2.04) | 0.43 | 1.04 | (0.62 - 1.76) | 0.74 |
|  | VA | 0.89 | (0.68 - 1.16) | 0.40 | 0.89 | (0.67 - 1.19) | 0.43 |
|  | CUIMC | 6.33 | (0.62 - 64.3) | 0.13 | 5.92 | (0.51 - 68.2) | 0.16 |
|  | OpenClaims | 1.08 | (0.91 - 1.28) | 0.38 | 1.05 | (0.90 - 1.24) | 0.53 |
|  | Optum DOD | 0.64 | (0.43 - 0.94) | 0.02 | 0.77 | (0.49 - 1.22) | 0.26 |
|  | Optum EHR | 1.21 | (0.40 - 3.69) | 0.74 | 1.36 | (0.33 - 5.66) | 0.67 |
|  | Meta-analysis | 0.98 | (0.85 - 1.14) | 0.83 | 1.00 | (0.89 - 1.13) | 0.94 |
| + Intensive services |  |  |  |  |  |  |  |
|  | SIDIAP | NA | NA | NA | NA | NA | NA |
|  | VA | 1.24 | (0.66 - 2.33) | 0.51 | 1.25 | (0.65 - 2.41) | 0.50 |
|  | CUIMC | NA | NA | NA | NA | NA | NA |
|  | OpenClaims | NA | NA | NA | NA | NA | NA |
|  | Optum DOD | 0.56 | (0.21 - 1.46) | 0.23 | 0.70 | (0.20 - 2.49) | 0.59 |
|  | Optum EHR | NA | NA | NA | NA | NA | NA |
|  | Meta-analysis | 1.16 | (0.74 - 1.80) | 0.52 | 1.15 | (0.71 - 1.88) | 0.56 |

**Figure 3.**
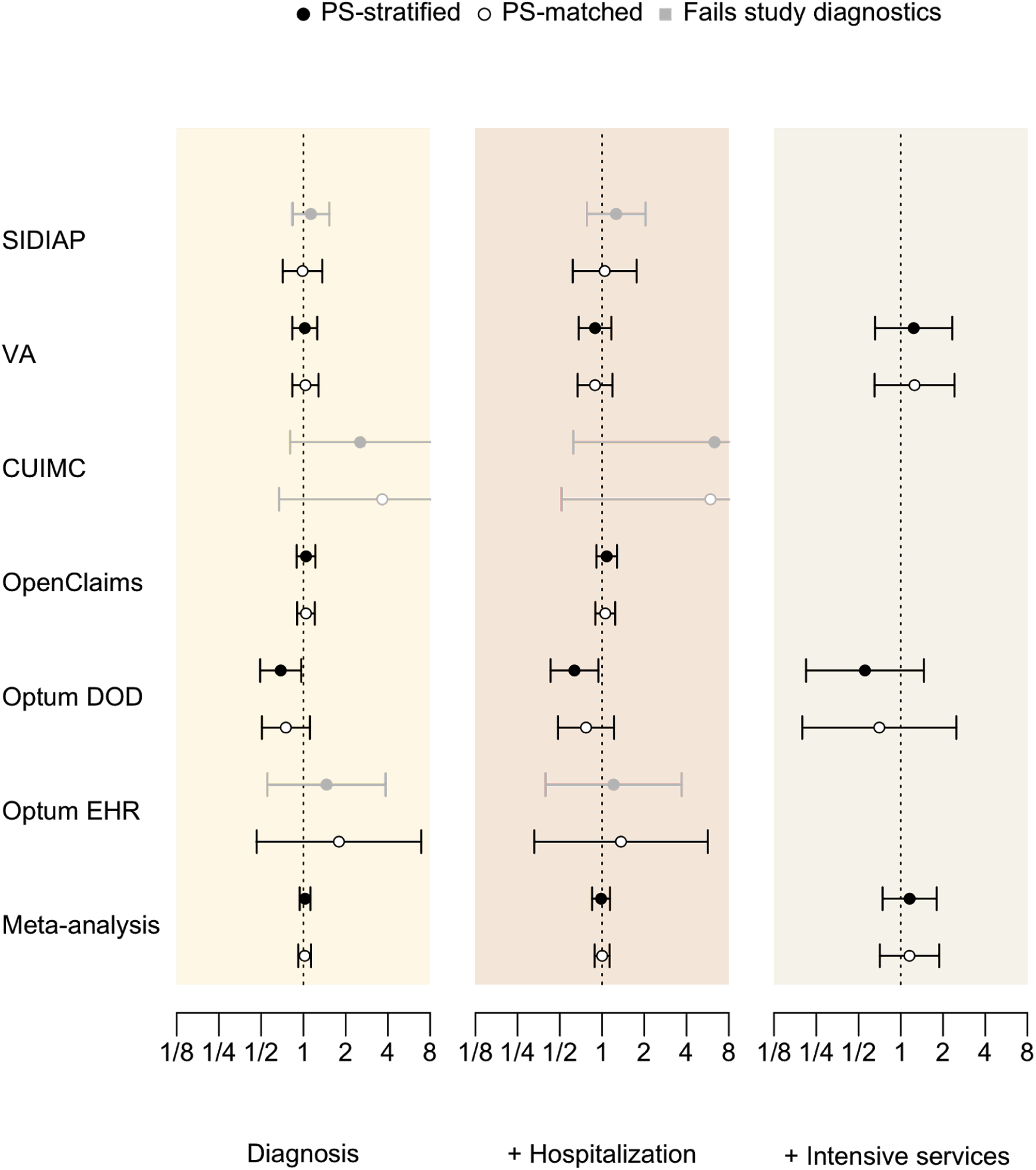
Hazard ratios of COVID-19 outcomes between alpha-1 blocker and 5ARI/PDE5 prevalent-use across data sources. The outcomes are COVID-19 diagnosis (Diagnosis), COVID-19 hospitalization (+ Hospitalization), and COVID-19 hospitalization requiring intensive services (+ Intensive services). We plot calibrated hazard ratios with black (PS-stratified) and white (PS-matched) circles along with their 95% confidence intervals. Grayed out entries do not pass study diagnostics.

For COVID-19 hospitalization, PS-matched analyses again found no differential risks according to drug use in any of the contributing data sources, with calibrated HRs of 1.04 (0.62 - 1.76) in SIDIAP, 0.89 (0.67 - 1.19) in VA, 1.05 (0.90 - 1.24) in OpenClaims, 0.77 (0.49 - 1.22) in Optum DOD, 1.36 (0.33 - 5.66) in Optum EHR, and 1.00 (0.89 - 1.13) in the meta-analysis.

For COVID-19 hospitalization requiring intensive services, only VA and Optum DOD passed diagnostics, with PS-matched calibrated HRs 1.25 (0.65 - 2.41) and 0.70 (0.20 - 2.49) respectively. Meta-analytic HR was 1.15 (0.71 - 1.88).

Out of the 118 pre-selected negative control outcomes, we used from 22 to 101 of them within each data source as we did not find sufficient numbers of events for the rest of negative controls.

## Discussion

In this international cohort study following more than 3.1 million men with BPH, we observed no association between alpha-1 blocker use and the risk of COVID-19 diagnosis, hospitalization, or hospitalization requiring intensive services. These findings bear out from 6 large real-world data sources, including out- and in-patient electronic medical records and health claims data from the US and Spain.

To our knowledge, Vogelstein et al. 2020 is the only existing epidemiological study exploring the potential association between alpha-1 blockers use and disease progression induced by lower respiratory tract infection. In this retrospective analysis of patients with acute respiratory distress or pneumonia, previous users of alpha-1 blockers — as compared to non-users — demonstrated lower risks of progression to ventilation and/or death. Given the very limited set of covariates considered in their analyses, however, there is serious concern for substantial residual confounding. In fact, with alpha-1 blockers being used to treat various diseases, confounding by indication may be severe enough that covariate adjustments cannot possibly account for systematic difference between the users and non-users. Moreover, healthy adherer bias (Hollestein et al. 2015) could have been introduced by their choice to include only patients who were prescribed alpha-1 blockers and had a medication possession ratio ≥ 50% in the year before the index date. Persistent alpha-1 blocker users likely have healthier lifestyles, such as sensible diet and regular exercise, than people who do not use or adhere to the medication. Such healthy adherer bias may have distorted their estimates towards the protective.

In our study, we selected 5ARI and PDE5 inhibitors as active comparators to minimize confounding by indication. We then confined our study population to patients with prior diagnosis of BPH in order to account for the fact that these drugs do not share all indications. In addition to the active comparator selection, we applied large-scale propensity score models involving tens of thousands of clinical covariates, thereby balancing a broad range of baseline patient characteristics. The conventional approach to adjust only for a small number of pre-selected covariates would have left many baseline characteristics imbalanced, which is particularly problematic when clinicians’ knowledge regarding this novel disease is limited.

The initial rationale that the disruption of catecholamine loop reduces CRS resulting from bacterial and nonbacterial causes was based on an animal study published in 2018 (Staedtke et al. 2018). After the pandemic began, the authors from this study postulated that prophylactic use of alpha-1 blockers might decrease the risk of progression to life-threatening complication among COVID-19 patients (Konig et al. 2020). This hypothesis was formed when three pieces of information were stacked: first, severe COVID-19 patients were often accompanied by a significant elevation of cytokines such as interleukin-6, interleukin-10 and tumour necrosis factor α. Second, catecholamines augmented the production of those cytokines in vitro and mice. Last, blockade of the alpha-1 adrenergic receptor (target of catecholamines) suppresses hyperinflammatory state in the context of bacterial infections.

However, we argue that preventing severe illness from the SARS-CoV-2 by targeting catecholamine-cytokine axis with the use of alpha-1 blockers is far more complicated and multifaceted. For example, use of alpha-1 blockers might increase the release of catecholamines through the negative feedback loop (Zuber et al. 2011), which in turn counteracts the benefit resulting from inhibition of alpha-1 adrenergic receptors. One the other hand, the surge of cytokines is more likely to be a consequence of patients responding over aggressively to an infection rather than a trigger. Therefore, the reduction of cytokines by deactivation of alpha-1 adrenergic receptors may not influence the CRS that already occurred.

### Strengths and limitations

This open science study comes with certain limitations, but also with unique strengths by virtue of our access to an international network of standardized databases. Below, we discuss potential limitations one by one, as well as ways through which our study addresses them to the extent practically possible.

First, the study only partially addresses the question of whether alpha-1 blockers alleviate the disease progression of COVID-19 as postulated by Konig et al. 2020. With COVID-19 being an emergent disease, the number of in-patient COVID-19 cases was rather small during the studied period even in the extensive network of databases we have access to. We thus determined the number to be insufficient for us to directly estimate the effectiveness against the disease progression in a scientifically meaningful manner. Instead, we attempted to investigate the question under the hypothesis that, if alpha-1 blockers were indeed protective against severe COVID-19 symptoms, we should see a negative association between the prevalent use of alpha-1 blockers and COVID-19 related outcomes such as hospitalization. Even COVID-19 diagnosis alone could be indicative of relatively severe symptoms since patients are otherwise unlikely to seek interactions with healthcare systems.

Second, we used a prevalent-user cohort design since there are so few patients initiating BPH therapies during and immediately preceding the pandemic that a new-user design is infeasible. A prevalent-user design is susceptible to potential bias due to time-varying hazards and/or inclusion of treatment effect mediators in the adjustment. In particular, our finding does not eliminate the possibility that the incident use — but not prevalent use — of alpha-1 blockers protects against COVID-19.

Third, while we have taken great care in identifying prescriptions of the study drugs, patients’ adherence to prescribed drugs cannot be determined from secondary observational health data. Presence (or absence) of prescription records in EHR or claims databases does not guarantee that the patient was in fact taking (or not taking) the prescribed drug. We mitigate this problem, however, by applying an active comparator for which we expect the misclassification rate to be similar to the target drug. This way, uncertainty in the drug usages would affect the magnitude of hazard ratios, but not its direction.

Fourth, another important limitation is under-diagnosis or under-reporting of COVID-19. We alleviated this issue by using a definition of COVID-19 based on broad data sources, including clinical diagnosis and/or PCR test data. However, this strategy still cannot account for many infected patients who likely remain asymptomatic or do not seek health care services. The issue is further exacerbated by the fact that the diagnosis and reporting of COVID-19 related data vary significantly over different sites and at different time points in the pandemic. While no observational study of a potential COVID-19 treatment is immune to these caveats, we believe that our approach — with consistent applications of the same design and analysis over an international network of observational databases — provides some of the most reliable real-world evidence. We found some variation in the estimates across our databases but not at a statistically significant level, giving us confidence in our study design choice and overall conclusion. Single-center observational studies, on the other hand, have no ways to assess whether their findings would hold under different data sources.

Finally, we conducted this study among adult men with BPH to guard against confounding by indication. Thus, these findings may not generalize to wider populations. However, older male patients constitute a particularly relevant high-risk subpopulation, accounting for a substantial portion of the severe cases of COVID-19. Also, there is currently no evidence that the pathophysiology of BPH modifies the effect of alpha-1 blockers on COVID-19.

### Clinical implications and conclusions

Among male patients with BHP, our findings do not support prophylactic use of alpha-1 blockers to decrease their risk of COVID-19 infection and progression. Further research is needed to determine the potential therapeutic effect of alpha-1 blocker initiation on people who are recently infected with SARS-CoV-2.

## Methods

### Study design

We conducted a prevalent-user active comparator cohort analysis across an internationally distributed network of databases. Our protocol is available at https://github.com/ohdsi-studies/Covid19SusceptibilityAlphaBlockers and registered in the EU PAS register (EUPAS36231).

The study design described in this section is visually summarized in a schematic diagram of Supplementary Figure 1 in the appendix.

### Data sources

We obtained routinely-collected electronic health records (EHRs) and claims data from Spain and the United States (US). All data sources had been previously mapped to the Observational Medical Outcomes Partnership (OMOP) Common Data Model (CDMv5) (Hripcsak et al. 2015). This enabled distributed network analyses without sharing patient-level data, whilst ensuring data provenance, by applying common analytical programmes across data partner centers. The included data sources are:

- Information System for Research in Primary Care (SIDIAP) database, covering approximately 80% of the population of Catalonia, Spain, or six million patients in number. SIDIAP contains data since 2006 from general practice EHRs linked to hospital admissions with information on diagnoses, prescriptions, laboratory tests, and lifestyle and sociodemographics and the central database of RT-PCR COVID-19 tests;
- US Department of Veterans Affairs (VA) database, covering approximately 12 million patients from 170 medical centers across the US and including administrative, clinical, laboratory, and pharmacy data repositories that are linked using unique patient identifiers (Lynch et al. 2019);
- Columbia University Irving Medical Center data warehouse (CUIMC) EHRs covering approximately six million patients from the New York-Presbyterian Hospital/Columbia University Irving Medical Center in the US. CUIMC includes data on clinical diagnoses, prescriptions, laboratory tests, demographics, and diagnosis and test for COVID-19;
- IQVIA Open Claims, covering approximately 160 million patients in the US and providing pre-adjudicated health insurance claims at the anonymized patient-level, collected from office-based physicians and specialists via office management software and clearinghouse switch sources for the purpose of reimbursement;
- Optum^®^ De-Identified Clinformatics Data Mart Database — Date of Death (Optum DOD), covering approximately 86.8 million patients under private health insurance mostly in commercial plans but also in Medicare Advantage. Optum DOD includes data captured from administrative claims processed from inpatient and outpatient medical services and prescriptions as dispensed, as well as results for outpatient lab tests processed by large national lab vendors who participate in data exchange with Optum;
- Optum^®^ De-Identified COVID-19 Electronic Health Record (Optum EHR), covering approximately 1.73 million patients from a network of healthcare provider organizations across the US. Optum EHR includes data since 2007 on demographics, medications prescribed and administered, lab results, vital signs and other observable measurements, clinical and inpatient stay administrative data, diagnosis and test for COVID-19.

Each site obtained institutional review board approval, or confirmed the study to be exempt or deemed not human subjects research. At the time of this study, SIDIAP and IQVIA were last updated in July, 2020; CUIMC in August, 2020; and VA, Optum DOD, and Optum EHR in September, 2020.

### Cohort eligibility, study period and follow-up

Each cohort consisted of adult males aged 18 years or older who received at least one eligible prescription or dispensation for one of the study drugs between November 1st, 2019 and January 31st, 2020. Index date was set as the date of the last prescription in this time window. We required participants to be observable in their data source for at least 180 days prior to the index date and to have a recorded history of BPH at any time prior to or on the index date (Supplementary Figure 1). Participants were then followed up until the earliest of: occurrence of an outcome; end of exposure; death; loss or deregistration from the database; or date of last data collection.

### Exposures

We compared exposures to alpha-1 blockers with exposures to other drug classes commonly indicated for treatment of BPH as active comparators. More precisely, the comparator consisted of dutasteride and finasteride (5ARI) and tadalafil (PDE5 inhibitor). The OMOP CDM concept IDs for these drug classes are provided in the supplement.

We restricted our analysis to subjects under monotherapy at cohort entry, excluding those who were exposed to alternative BPH treatments any time within 180 days prior to and including the index date. With the exception of our analysis on the Optum databases, we defined continuous drug exposures from the start of follow-up by grouping sequential prescriptions that have ≤ 30-day refill gaps between them. The end of exposure was defined as the end of the last prescription’s drug supply in such a sequence. In the Optum databases, it is difficult to identify periods of continuous drug exposure as prescriptions are not recorded consistently. We therefore used an intent-to-treat (ITT) type analysis, following patients until their record ends regardless of treatment persistence.

### Outcomes

We investigated three outcomes: 1) COVID-19 diagnosis; 2) COVID-19 hospitalization (inpatient visit with COVID-19 diagnosis during or up to three weeks prior to hospitalization); 3) COVID-19 hospitalization with intensive services (mechanical ventilation, tracheostomy, or extracorporeal membrane oxygenation). Positive tests results or diagnostic codes defined COVID-19 status. We provide the full details of our cohort and outcome definitions in the supplement, as well as in the online protocol.

### Study size

We included all patients meeting the eligibility criteria within each database and hence performed no a priori sample size calculation. Instead, we provided a minimum detectable rate ratio (MDRR) for each target-comparator-outcome triplet across each data source. MDRR is for achieving 5% type-1 error rate and 80% power.

### Statistical analyses

To adjust for measured confounding and improve covariate balance between comparison cohorts in a data-driven manner, we built large-scale PS models and fit them via regularized regression (Rosenbaum and Rubin 1983; Tian, Schuemie, and Suchard 2018). Our large-scale PS models consist of a broad range of pre-defined clinical covariates — including age, gender, race (US data) and other demographics, prior conditions, drug exposures, procedures, and health service utilization behaviors — to allow for the most accurate prediction of treatment and balance cohorts across many characteristics. For computational efficiency, we excluded all features that occurred in fewer than 0.1% of patients within the target and comparator cohorts prior to PS model fitting.

In separate analyses, we stratified into 5 PS quintiles or variable-ratio matched patients by PS, and used Cox proportional hazards models to estimate hazard ratios (HRs) between alternative target and comparator treatments for the risk of each outcome in each data source. The regression conditioned on the PS strata/matching-unit with treatment allocation as the sole explanatory variable. We aggregated HR estimates across data sources to produce meta-analytic estimates using a random-effects meta-analysis with inverse-variance weights (DerSimonian and Laird 1986). To study three outcomes in six data sources (plus one meta-analysis) using two PS-adjustment approaches, we generated 3 x (6 + 1) x 2 = 42 study effects.

Residual bias often remains in observational studies even after controlling for measured confounding through PS-adjustment (Schuemie et al. 2014, 2016). For each study, therefore, we conducted negative control outcome experiments, where the null hypothesis of no effect is believed to be true, using up to 118 controls identified through a data-rich algorithm (Voss et al. 2017) and then reviewed by clinicians. Using the empirical null distributions from these experiments, we calibrated each study effect HR estimate, its 95% confidence interval (CI) and the *p*-value to reject the null hypothesis of no differential effect (Schuemie et al. 2018). We declared a HR as significantly different from no effect when the calibrated *p*-value is less than 0.05, without correcting for multiple testing.

Blinded to the results, clinicians and epidemiologists evaluated study diagnostics for these treatment comparisons to assess if they were likely to yield unbiased estimates. The suite of diagnostics included (1) MDRR, (2) distributions of preference score, a transformation of PS that accounts for difference in exposure prevalences (Walker et al. 2013), to evaluate empirical equipoise and population generalizability, (3) extensive patient characteristics to evaluate cohort balance before and after PS-adjustment, and (4) negative control calibration plots to assess residual bias. We defined target and comparator cohorts to stand in empirical equipoise if the majority of patients in both carry preference scores between 0.3 and 0.7 and to achieve sufficient balance if all after-adjustment baseline characteristics return absolute SMD < 0.1 (Austin 2009).

### Study execution

We conducted this study using the open-source OHDSI CohortMethod R package (https://ohdsi.github.io/CohortMethod/) with large-scale analytics made possible through the Cyclops R package (Suchard et al. 2013). Start-to-finish open and executable source code is available at: https://github.com/ohdsi-studies/Covid19SusceptibilityAlphaBlockers. To promote transparency and facilitate sharing and exploration of the complete result set, an interactive web application (https://data.ohdsi.org/Covid19SusceptibilityAlphaBlockers/) serves up study diagnostics and results for all study effects.

## Data Availability

"We obtained routinely-collected electronic health records (EHRs) and claims data from Spain and the United States (US). All data sources had been previously mapped to the Observational Medical Outcomes Partnership (OMOP) Common Data Model (CDMv5) (Hripcsak et al. 2015). This enabled distributed network analyses without sharing patient-level data, whilst ensuring data provenance, by applying common analytical programmes across data partner centers. Each site obtained institutional review board approval, or confirmed the study to be exempt or deemed not human subjects research."

## Acknowledgements

We acknowledge the tremendous work and dedication of the 350 participants from 30 nations in the March 2020 OHDSI COVID-19 Virtual Study-a-thon (https://www.ohdsi.org/covid-19-updates/), without whom this study could not have been realized. We would also like to acknowledge the healthcare professionals who collected the utilized data whilst caring for COVID-19 patients. Finally, we would like to remember the people who suffered and died from COVID-19 and their families and friends.

## Author contributions

AN, JX, DPA, and MAS were responsible for interpreting the results and writing the manuscript. AN and MAS were responsible for the study implementation and analysis. AN, KK, TDS, SFB, MA, CB, AS, SLD, KL, MEM, TF, DPA, and MAS (Data holders + AN & DPA & MAS) contributed to the study execution. AN, DRM, MMC, SCY, NP, DPA, and MAS (ICARIUS designer + AN & DPA & MAS) contributed to the study design. JW, AGS, MJS, JR, and PRR (Infrastructure) contributed to the study implementation. All the co-authors contributed to writing the manuscript. The corresponding author confirms that all authors read and approved the final manuscript.

## Ethical Approval

All data partners received IRB approval or waiver in accordance with their institutional governance guidelines. Use of the SIDIAP data source was approved by the Clinical Research Ethics Committee of IDIAPJGol (project code: 20/070-PCV). Use of the VA-OMOP data source was reviewed by the Department of Veterans Affairs Central Institutional Review Board (IRB) and was determined to meet the criteria for exemption under Exemption Category 4(3) and approved the request for Waiver of HIPAA Authorization. Use of the CUIMC data source was approved by the Columbia University IRB as an OHDSI network study (IRB# AAAO7805). IQVIA Open Claims, Optum EHR, and Optum DOD are commercially available, syndicated data assets that are licensed by contributing authors for observational research. These assets are de-identified commercially available data products that could be purchased and licensed by any researcher. The collection and de-identification of these data assets is a process that is commercial intellectual property and not privileged to the data licensees and the co-authors on this study. Licensees of these data have signed Data Use Agreements with the data vendors which detail the usage protocols for running retrospective research on these databases. All analyses performed in this study were in accordance with Data Use Agreement terms as specified by the data owners. As these data are deemed commercial assets, there is no Institutional Review Board applicable to the usage and dissemination of these result sets or required registration of the protocol with additional ethics oversight. Compliance with Data Use Agreement terms, which stipulate how these data can be used and for what purpose, is sufficient for the licensing commercial entities. Further inquiry related to the governance oversight of these assets can be made with the respective commercial entities: IQVIA (iqvia.com) and Optum (optum.com). At no point in the course of this study were the authors of this study exposed to identified patient-level data. All result sets represent aggregate, de-identified data that are represented at a minimum cell size of >5 to reduce potential for re-identification. Furthermore, the New England Institutional Review Board of Janssen Research & Development (Raritan, NJ) has determined that studies conducted on licensed copies of Optum databases are exempt from study-specific IRB review, as these studies do not qualify as human subjects research.

## Data Availability

We obtained routinely-collected electronic health records (EHRs) and claims data from Spain and the United States (US). All data sources had been previously mapped to the Observational Medical Outcomes Partnership (OMOP) Common Data Model (CDMv5) (Hripcsak et al. 2015). This enabled distributed network analyses without sharing patient-level data, whilst ensuring data provenance, by applying common analytical programmes across data partner centers.

## Supplementary Material

Available at: https://github.com/ohdsi-studies/Covid19SusceptibilityAlphaBlockers/blob/master/Documents/alpha_blocker_study_supplementary_result.pdf

